# An objective systematic comparison of the most common adverse events of COVID-19 vaccines

**DOI:** 10.1101/2021.10.11.21264830

**Authors:** C. Behrens, T. Gasperazzo, M. Samii-Moghadam, J.B. Lampe

**Affiliations:** Lampe & Company GmbH & Co. KG, Berlin, Germany

## Abstract

**Background:** Vaccination is an important tool in the fight against pandemics. However, the associated adverse events (AEs) may negatively impact the public perception of vaccines, therefore leading to decreased vaccination willingness. Interestingly, pharmacovigilance data of the three COVID-19 vaccines with a two-dose schedule approved in the EU (AstraZeneca, BioNTech and Moderna) already revealed country-specific differences in their safety profile early on (as of February 2021), at a time when the accumulated occurrence of specific AEs was not yet known. In the safety outcome assessment presented here, we aimed to establish whether these country-specific differences in pharmacovigilance data could be explained by differences in the frequency of AEs as reported in the respective approval studies of each vaccine.

**Methods:** A systematic search was performed to identify all publications regarding the randomized controlled trials (RCTs) of two-dose vaccines approved in the EU (AstraZeneca, BioNTech and Moderna), including regulatory reports and journal articles. All obtained safety data was manually entered into an SQL database. In order to enable the comparability among the data, the solicited AEs for all vaccines (i.e. those AEs actively sought after vaccination) were investigated. The data was standardized to promote comparability and overcome data heterogeneity and complexity.

**Findings:** Twelve documents regarding the RCTs for the three COVID-19 vaccines with a two-dose schedule approved in the EU (AstraZeneca, BioNTech and Moderna) were included in the safety outcome analysis. The entire safety data compiled in the SQL database amounted to 66 different study arms. The data structure revealed 13 different age thresholds or ranges and three different data sets regarding doses (first dose vs. second dose vs. all doses). After standardization and identification of subgroups, the analyses demonstrated that the highest rates of AEs occur after the first dose with the AstraZeneca vaccine, whereas with Moderna and BioNTech most AEs occur after the second dose. Astonishingly, the absolute frequencies of each AE after the first AstraZeneca dose correspond to those of the second dose of the mRNA vaccines (BioNTech and Moderna). Reversely, the absolute frequencies of the same AEs after the second AstraZeneca dose correspond to those of the first dose with the mRNA vaccines. The most common AEs with any vaccine were fatigue, headache and myalgia. Moreover, middle-aged subjects (18 to 55 years) had more side effects than older individuals (> 55 years), an observation that persisted among vaccines.

**Interpretation:** This is the first indirect comparison of these vaccines that uses all available RCT data. The absolute frequency of each AE is similar between the first AstraZeneca dose and the second dose of BioNTech or Moderna; their occurrence was thus independent of platform (vector or mRNA) or the vaccine itself. This assessment demonstrates that the varying frequencies of AEs reported in early pharmacovigilance data for the vaccines in distinct countries, at a time when the accumulated occurrence of specific AEs with certain vaccines was not yet known, cannot be explained by different frequencies being reported in the respective RCTs.

**Conclusion:** The approach presented here could help to objectify future discussions on vaccine preferences. Therefore, it may serve as basis for future public awareness campaigns and may also allow the comparison of vaccine performance in different subgroups (e.g. virus variants, high-risk patients). This approach may also be applied to a broad range of other challenges across the R&D process and various disease categories.

## Introduction

Vaccination is an important tool to combat viral infections. In the case of COVID-19, the approved vaccines are well tolerated and most adverse events (AEs) are only mild (WHO, 2021). However, unfavorable media reports about the associated AEs may negatively impact the public perception of certain vaccines and the vaccination willingness (Murphy et al., 2021).

The only objective sources of vaccine efficacy and safety data are randomized controlled trials (RCTs). These studies are essential to better estimate the relevance of more frequent AEs; however, more seldom AEs are not covered. In contrast, pharmacovigilance data is collected in larger cohorts after approval, allowing the detection of potentially very seldom AEs; therefore, it is believed to demonstrate a vaccine’s performance in the real world (Kesselheim et al., 2021). Nevertheless, pharmacovigilance data is collected in a non-randomized and uncontrolled way and may thus be subject to bias, reflecting the subjective perception of a vaccine’s safety in a population.

By now, the accumulated occurrence of specific AEs with certain vaccines is well known (PEI Sicherheitsbericht, 20 September 2021). Nevertheless, as regards overall reactogenicity, AstraZeneca already had a negative image in some countries right from the start (as of February 2021), as reflected in pharmacovigilance data indicating higher AE rates with this vector vaccine than with the available mRNA vaccines (BioNTech and Moderna) in Germany [7.6 in 1,000 vs. 1.6–2.9 per 1,000 vaccinations, respectively] (PEI Sicherheitsbericht, 26 February 2021) in contrast to the UK [3–4 in 1,000 vaccinations for both AstraZeneca and BioNTech vaccines] (MHRA Yellow Card Reporting, 5 February 2021). This is of importance, since a negative vaccine’s image might lead to reduced vaccination willingness, whereas a positive vaccine image might lead to an underreporting of AEs with substantial health risks. Considering all this, a comparison of the subjective pharmacovigilance data with the objective RCT data would be highly desirable.

It is well established that AstraZeneca presents most AEs after the first vaccine dose, whereas mRNA vaccines show them after the second dose. However, it is still unknown how comparable the level of AEs is among the vaccines, regardless of following the first or second dose.

The present exemplary analysis focused on the safety profile of most common AEs of the three two-dose vaccines currently approved in the EU (AstraZeneca, BioNTech and Moderna). This approach aims to objectify the current discussion on the safety of different vaccines and may contribute to mitigate the currently widespread vaccination hesitancy. Moreover, it serves as an outstanding example of how an indirect comparison among different clinical studies can be accomplished by means of systematic, standardized assessments of the biomedical literature that goes beyond traditional reviews.

## Material and Methods

A systematic search was performed to identify journal articles and regulatory documents regarding the RCTs of the COVID-19 vaccines with a two-dose schedule approved in the EU: the mRNA vaccines BNT162b2 (Comirnaty) by BioNTech/Pfizer and mRNA-1273 (Spikevax) by Moderna; and AZD1222/ChAdOx1 (Vaxzevria), developed by AstraZeneca/Oxford University. All publications available as of May 31, 2021 reporting results of the RCTs that served as basis for the regulatory approval of these vaccines, by the EMA and/or FDA, were considered and included in the assessment.

All published vaccine data (i.e. study design, demographics, safety outcomes) was systematically identified and compiled in a specialized, relational SQL database. The data was structured and standardized in order to reveal nearly identical subgroups across the different vaccine types. In order to render the data as comparable as possible, all solicited AEs (i.e. those AEs actively sought after vaccination) (Hervé et al., 2019) reported for each vaccine were identified and manually entered into the database. To overcome heterogeneity and promote comparability, the AEs were standardized according to the Medical Dictionary for Regulatory Activities (MedDRA version 22.1, English). This way, the actual frequency of AEs could be determined and compared among vaccines. Only comparable data was included in the assessment, according to pre-established criteria (Figure 1). The data was analyzed and visualized with the software Spotfire (version 10.10.0).

**Figure 1.**
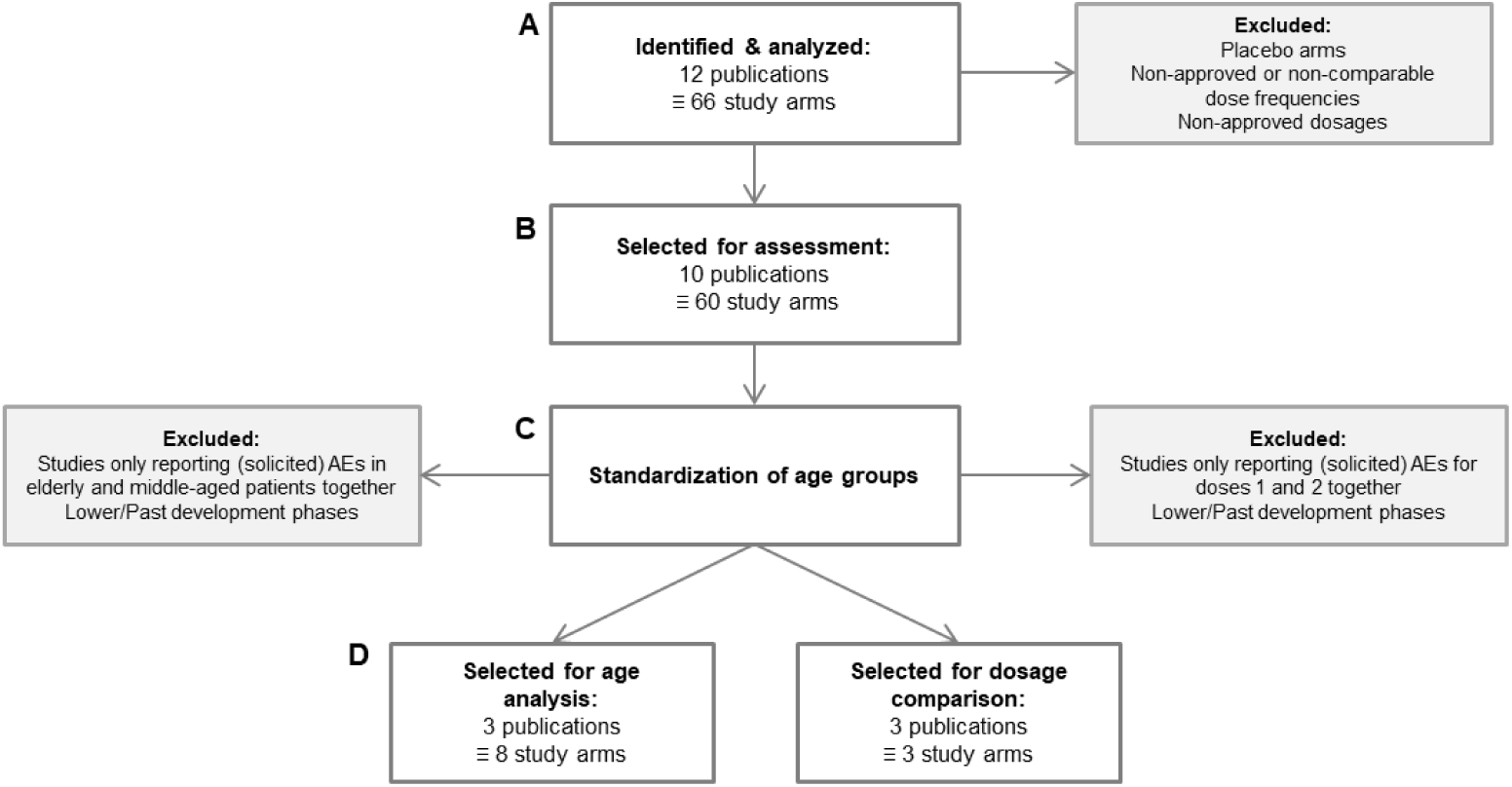
Flowchart showing the selection process for the data to be included in the present vaccine safety outcome assessment as described in Table 1. **A)** Identification of publications containing relevant information (i.e. prospective data from RCTs) about the safety of COVID-19 vaccines approved in the EU. **B)** Selection of the publications reporting approved and comparable vaccine dose frequencies and dosages. **C)** Standardization of the age groups identified in the publications selected in the previous step (Table 3). **D)** Selection of the publications with comparable data for subgroup analyses per age group as well as publications with safety outcomes reported separately for the first and second vaccine doses (Table 2).

## Results

Twelve publications reporting on RCTs were identified (Table 1), including ten journal articles and two regulatory documents (EMA). The entire data was compiled in the SQL database, amounting to 66 different study arms, corresponding to all information relevant for vaccine safety at the time of analysis. After exclusion of placebo arms and non-approved dosages and dosing schedules, ten publications were further assessed to identify comparable age groups (Figure 1). A standardization step revealed the suitability of three of the publications for subgroup analysis of vaccine AEs per age group and three publications for analysis per administration dose (Table 2): One EMA document (phase I–III) and one phase-II study were included for AstraZeneca, as well as one phase-III study each for BioNTech and Moderna, corresponding to three RCTs for the respective analyzed vaccines.

**Table 1.** Stepwise selection of publications (i.e. sources) containing relevant information to be included in the safety assessment of COVID-19 vaccines approved in the EU. Twelve publications were initially identified and compiled in a SQL database. Letters (A), (B), (C) and (D) correspond to the data selection steps presented in Figure 1. **A)** All twelve identified publications containing relevant information (i.e. prospective data from RCTs) about the safety of COVID-19 vaccines approved in the EU. **B)** Selected publications after exclusion of non-approved or non-comparable dose frequencies or dosages. **C)** Standardization of age groups in the selected publications. **D)** Selected publications with comparable safety outcomes reported per age group and for the first and second vaccine doses separately.

| Label | Trial Phase | ClinicalTrials.gov ID | Sources | (A) All | (B) Selected for Safety Assessment | (C) Standardization of Age Groups | (D) Selected for Subgroup Analysis per Age Group or Dose |
| --- | --- | --- | --- | --- | --- | --- | --- |
| AstraZeneca | Phase I/II/III | – | EMA Assessment Report: AstraZeneca (2021) | ✓ | ✓ | ✓ | ✓ |
|  | Phase I/II | NCT04324606 | Folegatti et al., 2020 | ✓ | ✓ | ✓ |  |
|  | Phase II/III | NCT04400838 | Ramasamy et al., 2020 | ✓ | ✓ | ✓ | ✓ |
|  |  |  | Voysey et al., 2020 | ✓ | ✓ | ✓ |  |
| BioNTech | Phase I/II | NCT04380701 | Sahin et al., 2020 | ✓ |  |  |  |
|  | Phase I/II/III | NCT04368728 | EMA Assessment Report: BioNTech (2021) | ✓ | ✓ | ✓ |  |
|  |  |  | Mulligan et al., 2020 | ✓ | ✓ | ✓ |  |
|  |  |  | Polack et al., 2020 | ✓ | ✓ | ✓ | ✓ |
|  |  |  | Walsh et al., 2020 | ✓ | ✓ | ✓ |  |
| Moderna | Phase I | NCT04283461 | Anderson et al., 2020 | ✓ | ✓ | ✓ |  |
|  |  |  | Jackson et al., 2020 | ✓ |  |  |  |
|  | Phase III | NCT04470427 | Baden et al., 2020 | ✓ | ✓ | ✓ | ✓ |

**Table 2.**
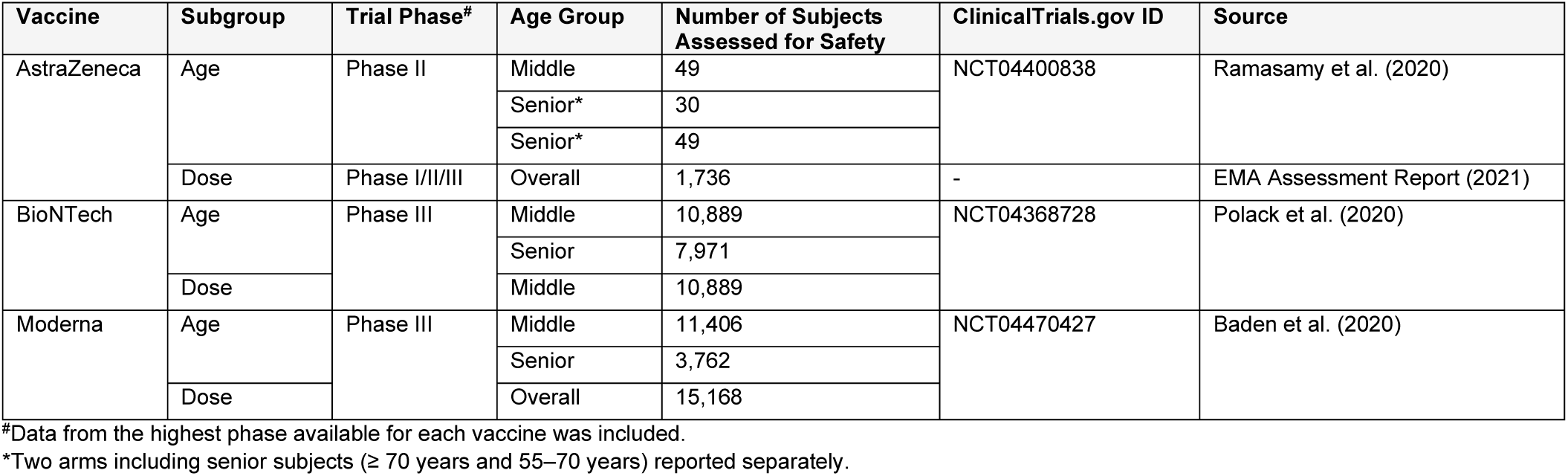
Details of the four publications selected for inclusion in the safety outcome assessment per subgroup (age and administration dose) after data standardization.

| Vaccine | Subgroup | Trial Phase<sup>#</sup> | Age Group | Number of Subjects Assessed for Safety | ClinicalTrials.gov ID | Source |
| --- | --- | --- | --- | --- | --- | --- |
| AstraZeneca | Age | Phase II | Middle | 49 | NCT04400838 | Ramasamy et al. (2020) |
|  |  |  | Senior* | 30 |  |  |
|  |  |  | Senior* | 49 |  |  |
|  | Dose | Phase I/II/III | Overall | 1,736 | - | EMA Assessment Report (2021) |
| BioNTech | Age | Phase III | Middle | 10,889 | NCT04368728 | Polack et al. (2020) |
|  |  |  | Senior | 7,971 |  |  |
|  | Dose |  | Middle | 10,889 |  |  |
| Moderna | Age | Phase III | Middle | 11,406 | NCT04470427 | Baden et al. (2020) |
|  |  |  | Senior | 3,762 |  |  |
|  | Dose |  | Overall | 15,168 |  |  |

<sup>#</sup>Data from the highest phase available for each vaccine was included.
\*Two arms including senior subjects (≥ 70 years and 55–70 years) reported separately.

**Table 3.** Age groups as reported in journal articles and regulatory documents and their standardization for analysis.

| Age as reported [years] | Standardized age groups |
| --- | --- |
| 16–55 | Middle |
| 18–55 |  |
| 18–59 |  |
| 18–64 |  |
| 16–89 | Overall |
| 16–91 |  |
| 18–95 |  |
| ≥ 18 |  |
| 55–70 | Senior |
| > 55 |  |
| ≥ 60 |  |
| ≥ 65 |  |
| ≥ 70 |  |

The age groups were standardized as shown in Table 3. Altogether, across the three analyzed vaccines, the data structure revealed 13 different age ranges (Table 3), three trial phases (I–III), three distinct dose analysis sets (first dose vs. second dose vs. all doses), and various numbers of subjects, depending on trial phase (Table 2).

The most common AEs with any vaccine were fatigue, headache and myalgia. Analysis of the data after standardization demonstrated that the AEs observed with highest frequency with the AstraZeneca vaccine occur after the first dose. Contrarily, with the Moderna and BioNTech vaccines the AEs observed with greatest frequency occur after the second dose (Figure 2). Moreover, the absolute frequencies of each AE after the first AstraZeneca dose correspond to those of the second dose of the mRNA vaccines (BioNTech and Moderna); reversely, the absolute frequencies of the same AEs after the second AstraZeneca dose correspond to those of the first dose with the mRNA vaccines (Figure 3). Overall, regardless of dose, the frequency of each AE was very similar across vaccines and did not show a tendency towards one vaccine over another, independent of the used platform (non-replicating viral vector or mRNA).

**Figure 2.**
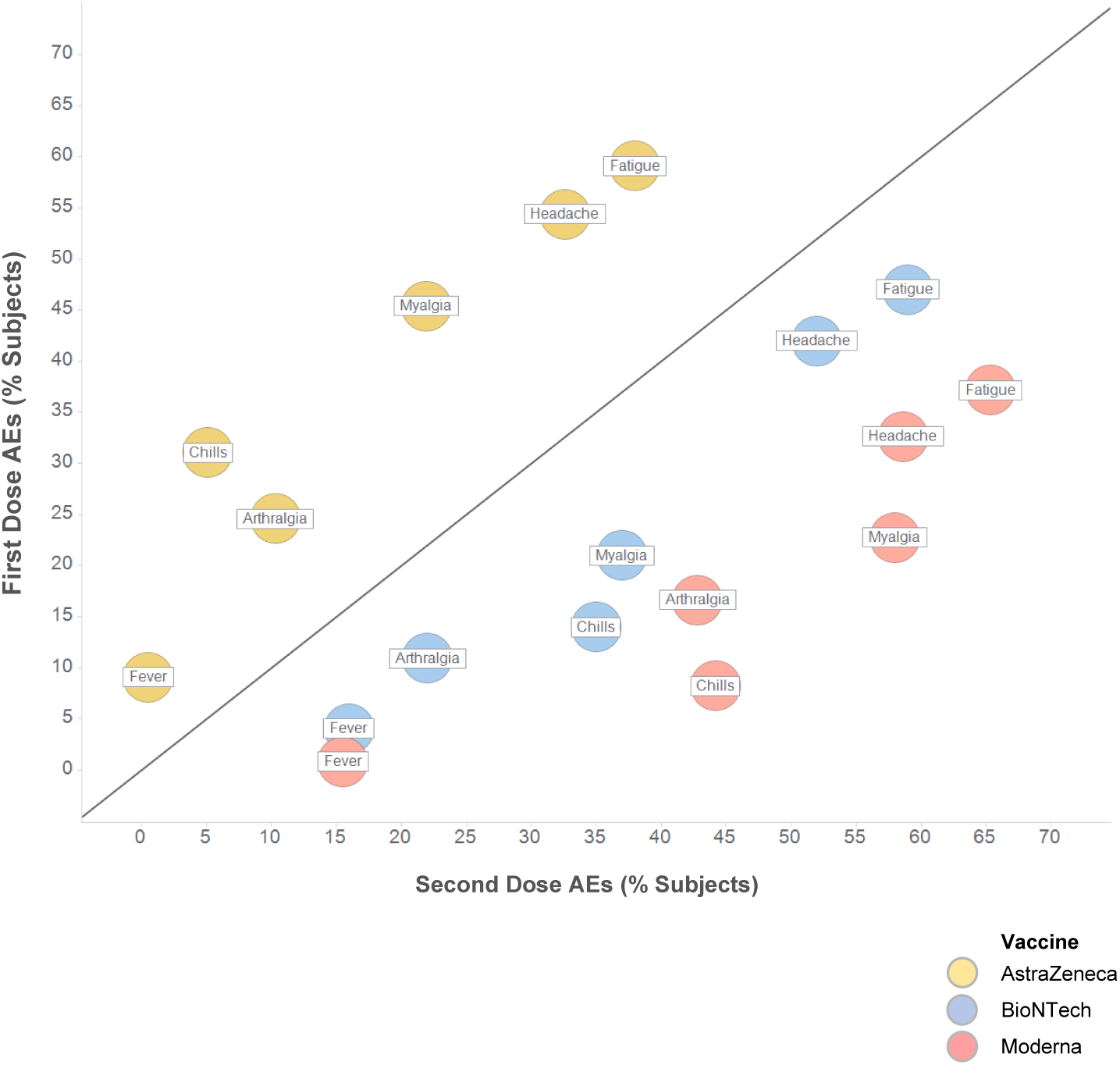
Most common systemic adverse events occurring after the first and second doses of anti-SARS-CoV-2 vaccines. The highest frequencies for each adverse event occurring with each vaccine are shown. For BioNTech only young/middle-aged subjects (18–65 years) are included; no data was available for overall population. Sources: AstraZeneca (AZ): EMA Assessment Report (2021); BioNTech (BNT): Polack et al. (2020); Moderna (MOD): Baden et al. (2020).

**Figure 3.**
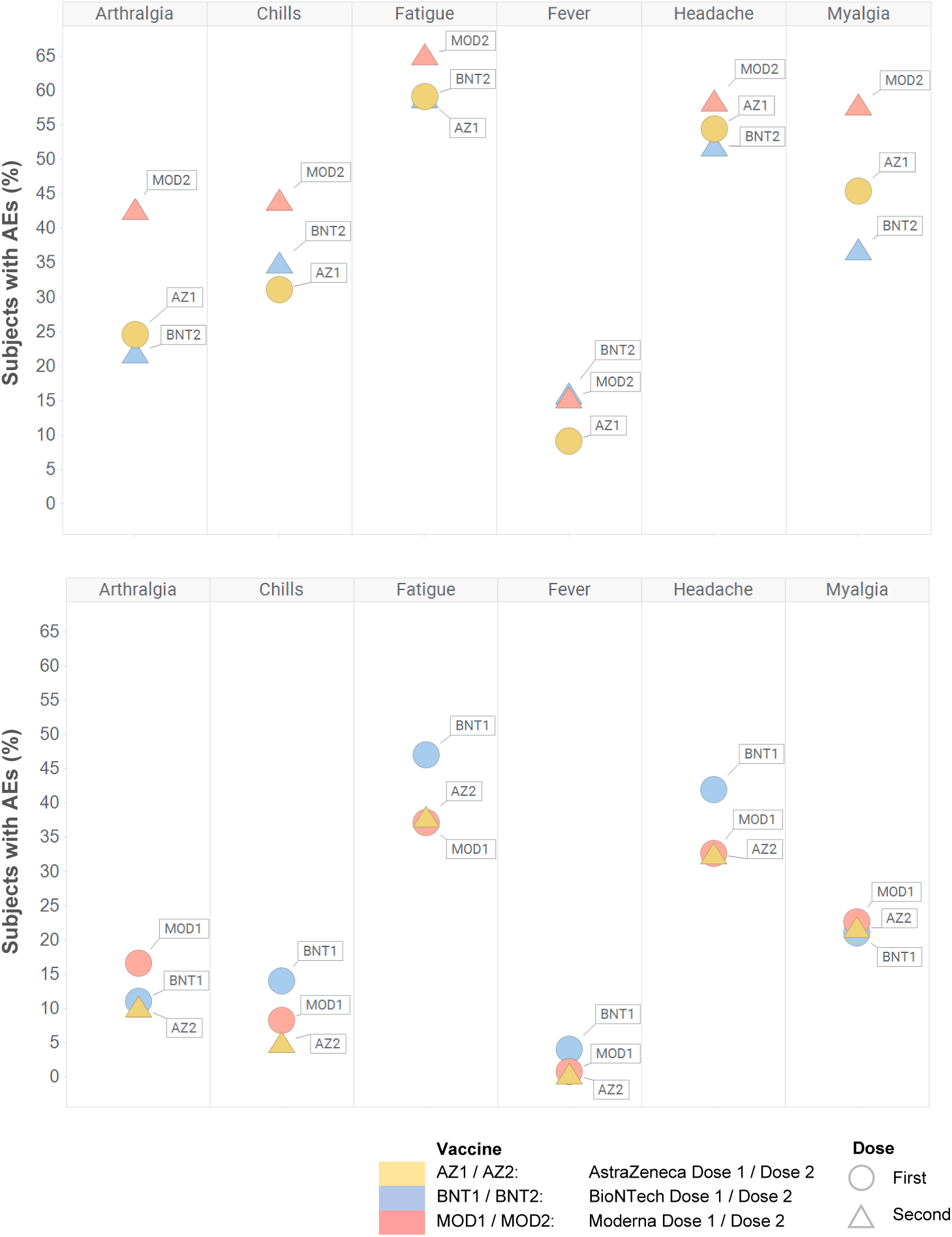
Plot showing that adverse event frequencies with the first dose of AstraZeneca (a viral vector vaccine) are similar to those with the second dose of BioNTech and Moderna (mRNA vaccines), and vice-versa. The highest frequencies for each adverse event occurring with each vaccine at each dose are shown. The numbers 1 and 2 after the vaccine names indicate the vaccine dose. For BioNTech only young/middle-aged subjects (18–65 years) are included; no data was available for overall population.

Further, middle-aged subjects (18 to 55 years) had more side effects than older individuals (> 55 years), an observation that remained constant among vaccines (Figure 4). Fatigue, headache and myalgia were mild in these subgroups, and prevailed as the most common AEs independent of age. There were no large differences among the vaccines for this age subgroup; however, for AstraZeneca, comparable data was only available from Phase I/II studies with few patients (n = 49), whereas BioNTech and Moderna studies included large cohorts (over 10,000 subjects each).

**Figure 4.**
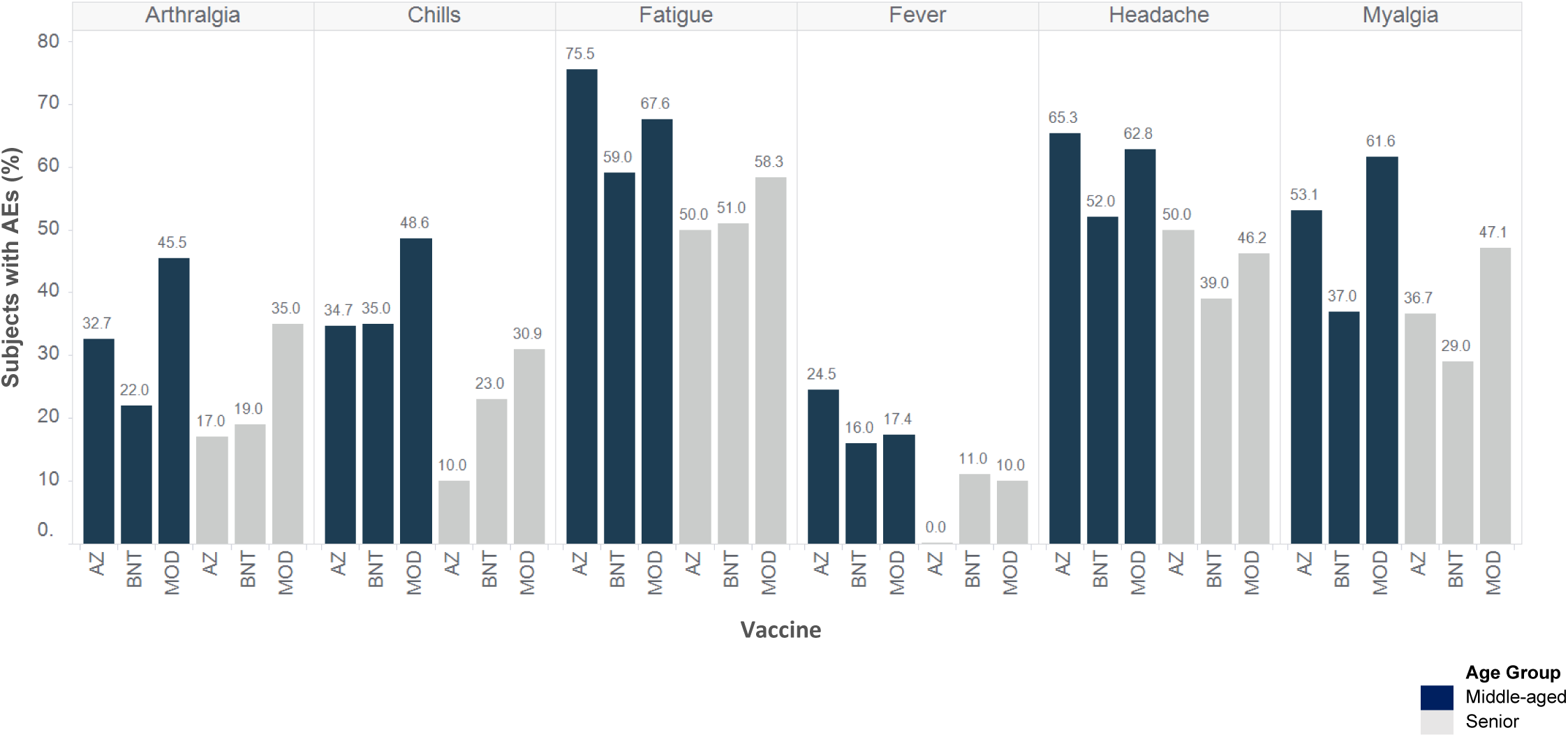
Overall frequency of systemic adverse events by age group observed with the anti-SARS-CoV-2 vaccines. Shown here are the highest observed frequencies of each adverse event, independent of dose (first or second). ^*^Maximum age for middle-aged subjects and minimum age for senior subjects varies from 55–65 years. Sources: AstraZeneca (AZ): Ramasamy et al. (2020); BioNTech (BNT): Polack et al. (2020); Moderna (MOD): Baden et al. (2020)

Unsurprisingly, blood clots and thrombosis were not reported in the included RCTs, which is likely due to their very rare occurrence in the population.

## Discussion

Comparisons between different studies may have a great impact on both individual treatment decisions as well as on decisions during the pharma R&D process. However, head-to-head comparisons are rarely conducted; instead, indirect comparisons are performed (e.g. Cochrane analyses, review articles).

Here we present a tried-and-tested approach to accomplish an indirect comparison of trial data, exemplarily applied to the three anti-SARS-CoV-2 vaccines with two doses currently approved in the EU. In order to bring the data to a comparable basis, all solicited AEs (i.e. those AEs actively sought after vaccination) (Hervé et al., 2019) reported for each vaccine were investigated.

Overall, the approved vaccines are well tolerated, and most adverse events (AEs) are only mild. Middle-aged subjects (18 to 55 years) had more side effects than older individuals (> 55 years), an observation that remained constant among vaccines. Fatigue, headache and myalgia were mild in these subgroups, and prevailed as the most common AEs independent of age.

Analysis of the standardized data on RCTs demonstrated that most AEs occur after the first dose with the AstraZeneca vaccine, whereas with Moderna and BioNTech most AEs occur after the second dose. Interestingly, the absolute frequencies of each AE after the first AstraZeneca dose correspond to those of the second dose of the mRNA vaccines (BioNTech and Moderna). Reversely, the absolute frequencies of the same AEs after the second AstraZeneca dose correspond to those of the first dose with the mRNA vaccines.

The accumulated occurrence of specific AEs with certain vaccines in distinct countries early on, at a time when these AEs were not yet well established, cannot be explained by the objective data derived from RCTs. This is of importance since a negative reporting bias of AEs might lead to reduced vaccination willingness, whereas positive bias might lead to an underreporting of AEs with substantial health risks.

The analyses presented here are a selection of the main results of a systematic safety assessment based on all available RCT data. To overcome the heterogeneity and complexity of the available information, the data has been sorted, categorized, standardized, analyzed, and finally visualized. The same approach could also be applied to evaluate other key vaccine features, such as efficacy regarding different viral variants. As a consequence, key messages on each vaccine could be immediately used for patient education, individual vaccine decisions or planning of future vaccination campaigns. This approach objectifies the current discussion on the safety of different vaccines and may contribute to mitigate the currently widespread vaccination hesitancy. In addition, it may serve as a fast and reliable tool for early decision making in future pandemic situations.

This is the first indirect comparison of the selected vaccines that uses all available RCT data. Here we showed how to make clear statements, with the help of a standardized and systematic assessment of the medical literature, that would not be possible otherwise. The example of safety of vaccines against SARS-CoV-2 was chosen here because of its current relevance; however, this approach can also be applied to a broad range of challenges across the R&D process, and to a vast spectrum of diseases regardless of the studied treatment approach.

## Conclusion

The multidimensional assessment of published vaccine data presented here may serve as basis for a public awareness campaign to combat vaccine hesitancy by determining and explaining the phenomena which the public perceives intuitively.

## Data Availability

All data produced in the present work are contained in the manuscript.

